# Metabologenomic approach reveals intestinal environmental features associated with barley-induced glucose tolerance improvements in Japanese cohort: a randomized controlled trial

**DOI:** 10.1101/2021.04.29.21256299

**Authors:** Yuka Goto, Yuichiro Nishimoto, Shinnosuke Murakami, Tatsuhiro Nomaguchi, Yuka Mori, Masaki Ito, Ryohei Nakaguro, Toru Kudo, Tsubasa Matsuoka, Takuji Yamada, Toshiki Kobayashi, Shinji Fukuda

## Abstract

Consumption of barley has been known to exert beneficial effects on metabolic disorders; however, it has also been reported that there are inter-individual differences in these responses. Recent evidence has suggested that these individual differences are mediated by the gut microbiota. Therefore, in the present study, we aimed to understand the relationship between the intestinal environment, including gut microbiota, and metabolic disorders. A randomized controlled trial in Japanese subjects with 4-week consumption of barley or control food was conducted. In this study, we analyzed the intestinal environment, including microbiota and their metabolites, and blood parameters were assessed collectively. We found that microbial genera *Blautia* and *Agathobacter* belonging to Lachnospiraceae, and fecal metabolites such as azelate were increased 1.31-fold, 1.84-fold, and 1.48-fold after barley consumption, respectively. Furthermore, the subjects whose glucose tolerance were slightly impaired showed improvement in their glucose tolerance index following the barley consumption. Additionally, the analysis showed that the increase in the abundance of the *Anaerostipes* was correlated with the improvement in the glucose tolerance index. Our findings indicate that the effects of barley consumption for glucose tolerance are partly defined by the intestinal environment of consumers, providing a quantitative measurement of the dietary effect based on the intestinal environment.

## Introduction

Barley is one of the first domesticated grains and has been cultivated for about 10,000 years. Recent studies have revealed that barley has the potential to improve certain blood parameters such as glucose tolerance and total blood cholesterol (1, 2). As barley contains a high amount of β-glucan, a type of soluble fiber, its effect on these blood parameters has been attributed to its high viscosity. It has been reported that barley-derived β-glucan conjugates with sugars and lipids in the human gastrointestinal lumen to inhibit their digestion and absorption (3, 4). In addition, a previous study showed that barley consumption increases *Prevotella/Bacteroides* ratio in barley-consuming healthy Swedish subjects with improved glucose tolerance (5). These reports indicate that β-glucan affects blood glucose levels in two ways: directly by binding to sugars and indirectly through changes in the intestinal environment.

Despite these studies, the complete picture on the underlying relationship between barley intake and glucose intolerance is yet to be revealed, as many reports suggest that there are multiple confounding factors. First, genetic background affects the blood parameters. It has been reported that the primary symptom in the early stage of diabetes is different in populations from different backgrounds. For example, Caucasians show insulin resistance, whereas East Asians show a decrease in insulin secretion (6). Second, it has been suggested that the effect of barley is partly due to the functions of the intestinal microbiome (5); however, within the same species of bacteria, the existence of different strains unique to each country has been implied. It has also been proposed that the differences in these strains can possibly lead to differences in the phenotypes. For instance, various strains of *Prevotella copri* have been observed in multiple countries, and their functions are also dissimilar (7). Moreover, there have been no studies reporting the effect of barley on the intestinal environment and glucose tolerance index in Asian populations, such as the Japanese, who are known to have unique microbiomes (8).

As explained above, the effect of barley intake on the intestinal microbiome and metabolome profiles is still unclear. To elucidate the mechanisms through which barley improves the blood parameters through the intestinal environment in Japanese subjects, a randomized double-blind controlled trial was conducted. The intestinal environment was evaluated using a metabologenomics approach, which combines mass spectrometry-based metabolome and high-throughput sequencing-based microbiome analyses.

## Results

### The effect of barley intake on blood parameters

We conducted a randomized, double-blind controlled trial in 19 Japanese subjects (Fig 1). Baseline clinical characteristics were similar in primary and secondary outcomes including blood glucose area under the curve (AUC) and incremental AUC (iAUC), insulin AUC and iAUC, fasting blood glucose and insulin and stool frequency in both groups (Table S1). First, we statistically tested each primary and secondary outcome; however, there were no significant differences observed in any outcomes (Table 1). To investigate the beneficial effect of barley, parameters obtained from the clinical blood test, which was performed at six different time points, were evaluated through multiple testing (Friedman test followed by Nemenyi’s post-hoc test). There were significant differences in sodium, total cholesterol, and total protein levels (Fig 2, q < 0.10, False Discovery Rate (FDR) corrected). Next, the post-hoc test revealed a significant decrease at T3 (end of test food intervention period) in total protein and cholesterol levels compared to C1 and T1, respectively. For sodium, a significant decrease was observed at C3 and T2 compared to T1.

**Table 1.** Effect size and *p*-values of primary and secondary outcomes. *p*-value and effect size were calculated by paired t-test and paired Cohen’s d, respectively. Lower and upper represent 95% confidence interval of effect size.

|  | C2 vs T2 |  | C3 vs T3 |  |
| --- | --- | --- | --- | --- |
|  | effect size<br>(95% CI) | *p*-value | effect size<br>(95% CI) | *p*-value |
| Blood glucose AUC (0–120 min) | -0.019<br>(-0.677 to 0.639) | 0.934 | 0.280<br>(-0.381 to 0.941) | 0.238 |
| Insulin AUC (0–120 min) | 0.018<br>(-0.640 to 0.676) | 0.937 | -0.128<br>(-0.787 to 0.530) | 0.582 |
| Blood glucose iAUC (0–120 min) | -0.104<br>(-0.762 to 0.555) | 0.657 | 0.269<br>(-0.392 to 0.930) | 0.256 |
| Insulin iAUC (0–120 min) | 0.018<br>(-0.640 to 0.677) | 0.937 | -0.095<br>(-0.754 to 0.563) | 0.682 |
| Fasting blood glucose | 0.252<br>(-0.409 to 0.912) | 0.287 | 0.022<br>(-0.636 to 0.680) | 0.924 |
| Fasting blood insulin | 0.009<br>(-0.649 to 0.667) | 0.968 | -0.289<br>(-0.951 to 0.372) | 0.223 |
| Stool frequency | 0.127<br>(-0.531 to 0.786) | 0.585 | 0.279<br>(-0.382 to 0.940) | 0.240 |

**Fig 1.**
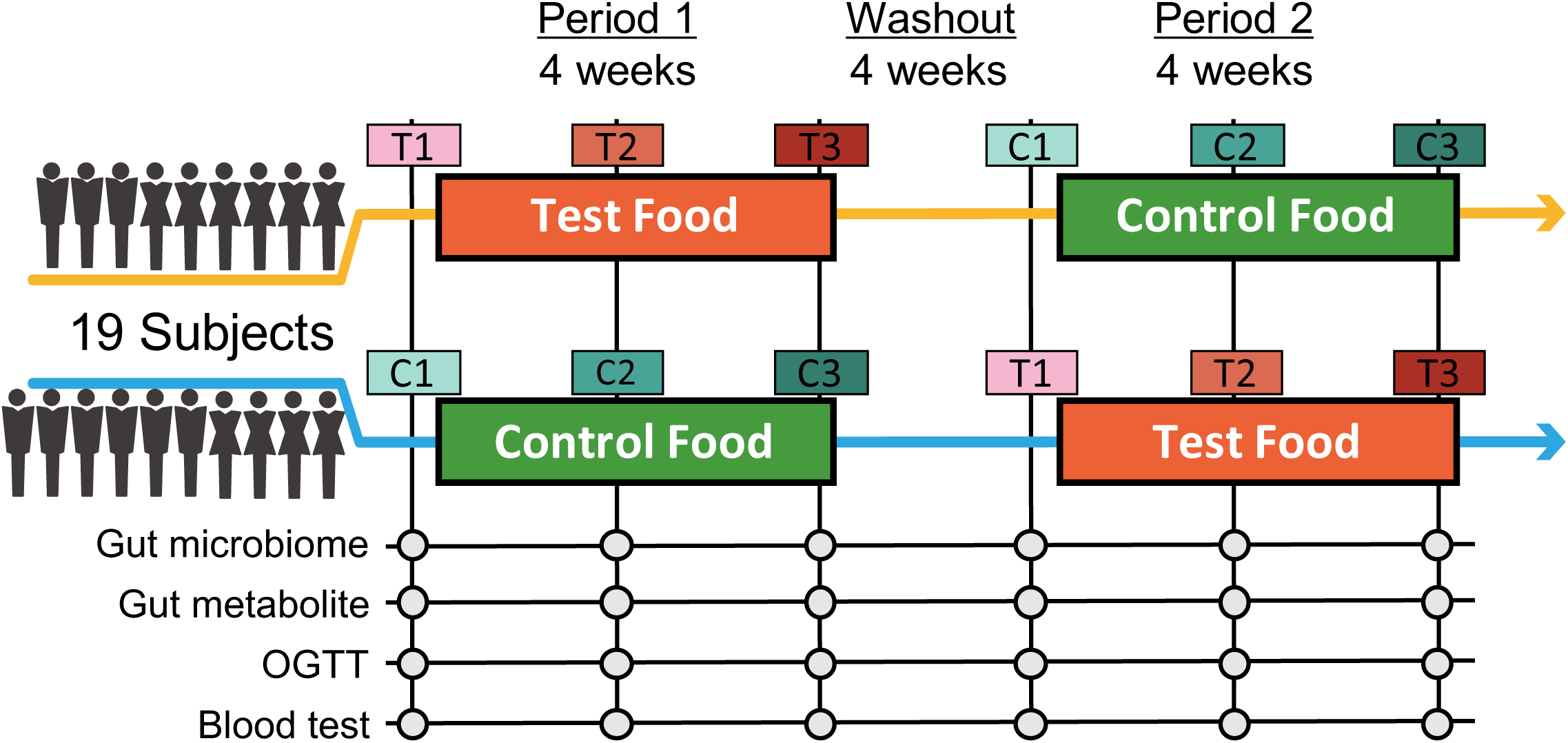
Timeline of the double-blind randomized crossover trial. Two four-week dietary treatments were set in succession. The dietary intervention periods were interspaced by a four-week washout period. Blood and stool samples were collected before and after 2 and 4 weeks of each intervention period.

**Fig 2.**
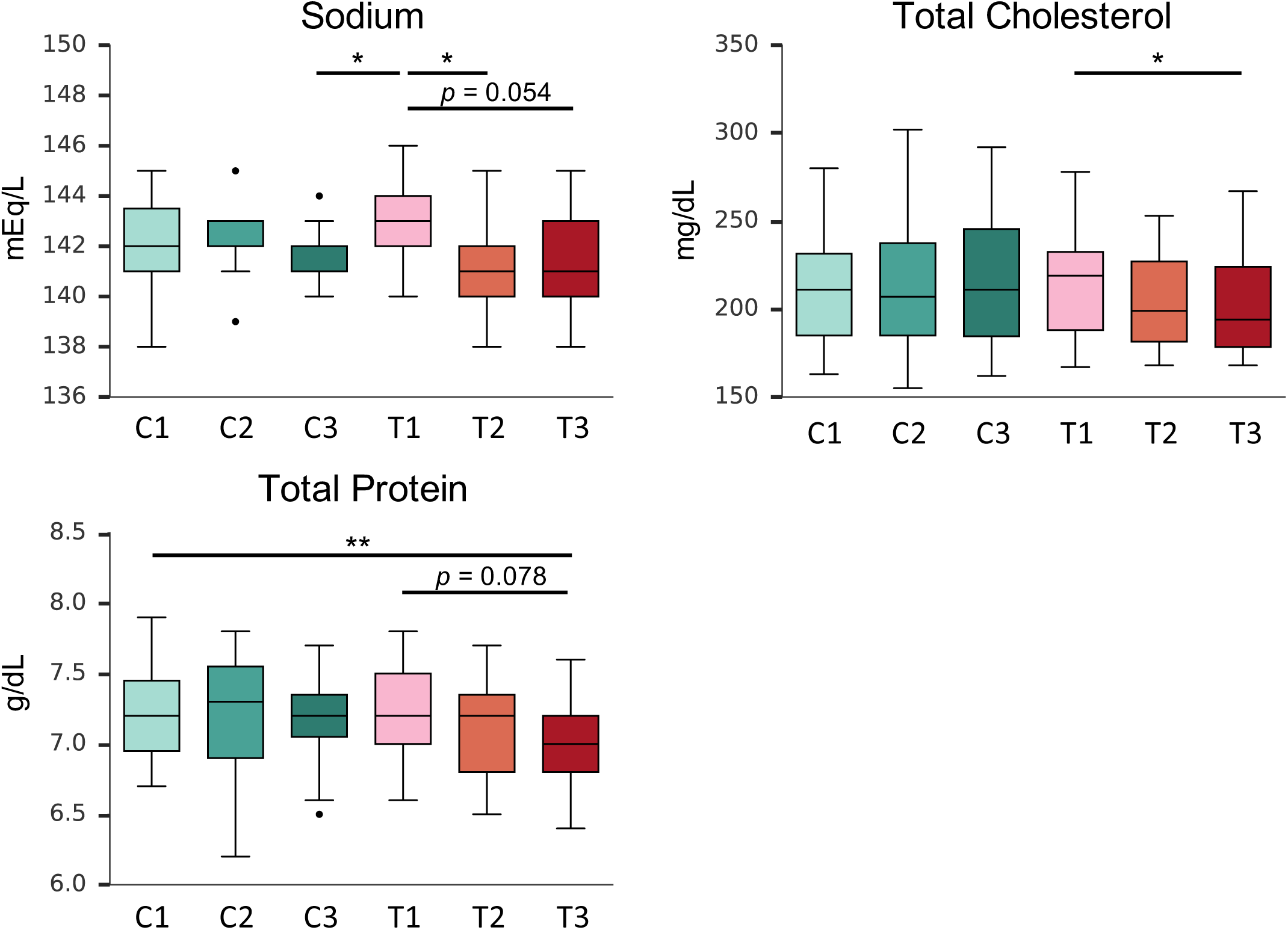
Barely consumption altered blood parameters significantly in this trial. Box plots showing significant differences in distribution of blood parameters at multiple time points (q < 0.10; Friedman test). Post-hoc tests were subsequently performed to identify time points showing significant differences (*, *p* < 0.05; **, *p* < 0.005; Nemenyi’s test). Labeled letters represent the time point of blood sample collection: T, intervention period with test food; C, intervention period with control food; T1/C1, before intervention; T2/C2, after 2 weeks of intervention; T3/C3, after 4 weeks of intervention.

### Effect of barley intake on intestinal microbiome and metabolome profiles

To evaluate the effect of barley on intestinal microbiome and metabolome profiles, we performed 16S rRNA gene-based microbiome analysis and CE-TOFMS-based metabolome analysis. Multidimensional scaling with beta diversity showed that the plots from each subject were clustered in both microbiome and metabolome profiles (Fig 3). Comparison of inter-individual distances and intra-individual distances showed significant differences (Wilcoxon rank sum test, *p* = 9.90*10^−180^ and *p* = 3.99 * 10^−116^ at microbiome and metabolome profile, respectively), suggesting that the individual difference was larger than the influence of barley or control food consumption (Fig 3C, F, Fig S1).

**Fig 3.**
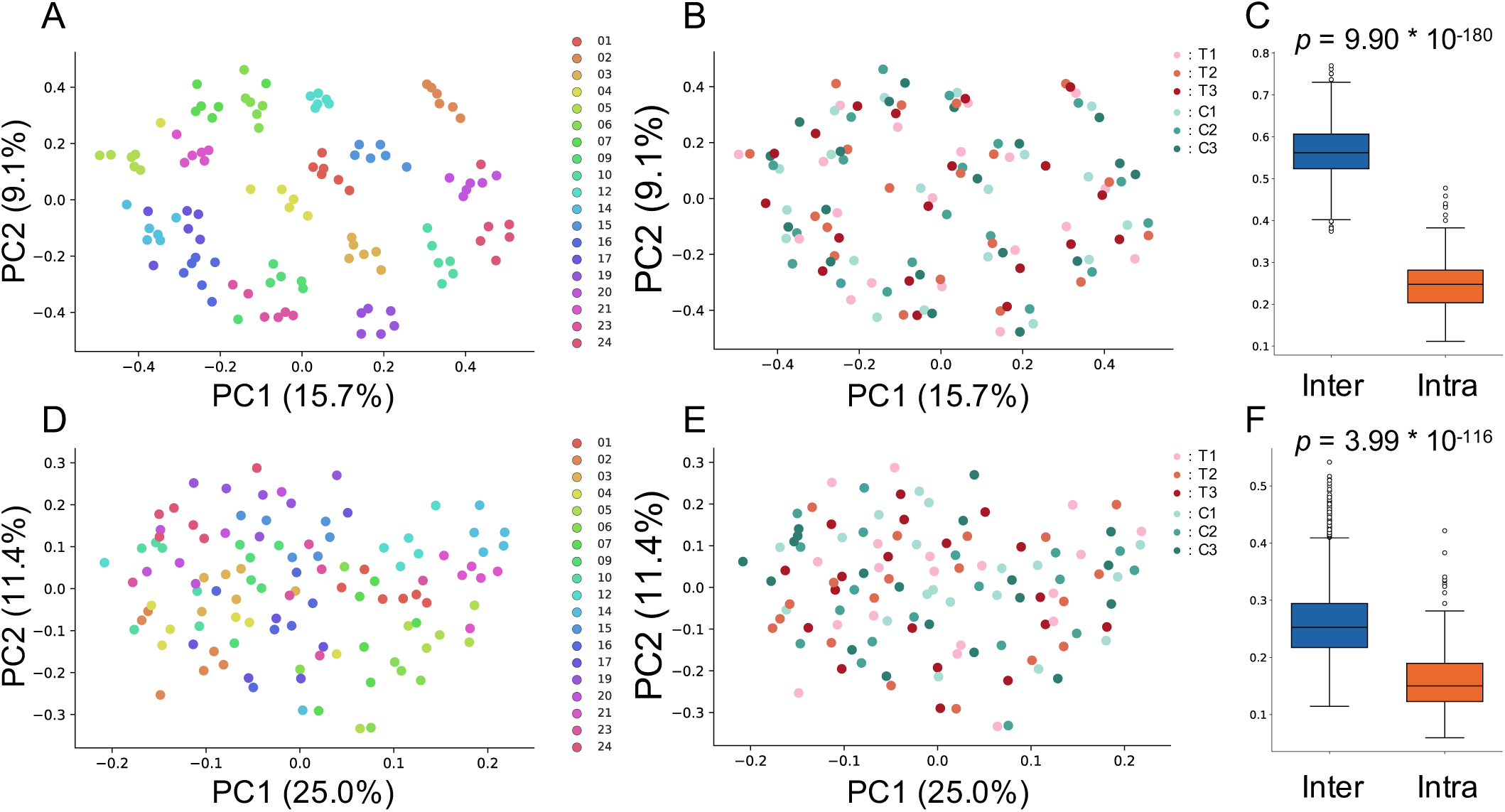
Individual differences were larger than the influence of barley or control food consumption on microbiome and metabolome profiles. (A,B) Scatter plots showing results of multidimensional scaling using beta diversity (unweighted UniFrac distance) calculated from microbiome profile. Plots were color-coded by subject (A) or time point (B). (C) Box plot representing the distribution of unweighted UniFrac distance between samples from different subjects at the same time point (inter-individual, represented in blue) and the distance between samples from the same subject (intra-individual, represented in orange). (D,E) Scatter plots showing the results of multidimensional scaling using beta diversity (Spearman correlation distance) calculated from metabolome profile. Plots were color-coded by subject (D) or time point (E). (F) Box plot representing distribution of Spearman correlation distance between samples from different subjects at the same time point (inter-individual, represented in blue) and the distance between samples from the same subject (intra-individual, represented in orange).

We next performed the Wilcoxon signed-rank test for the relative abundance of each microbe and the relative area of each metabolite, to further investigate the effect of barley consumption on the intestinal environment. The comparison was performed between pairs of two different sets; T1–T3 and C3–T3 (Fig 4). Some genera showed significant differences in their relative abundances (*p* < 0.05, not corrected), such as *Blautia* and those belonging to the family Lachnospiraceae in T1–T3 comparison; however, these differences were only observed as significant without multiple testing correction (Table S2). Additionally, several metabolites showed significant increases (*e*.*g*. azelate and imidazole propionate) and decreases (*e*.*g*. paraxanthine and 6-hydroxynicotic acid) in their relative area; however, the significance was only observed without multiple testing correction (Fig 4C, D; Table S3).

**Fig 4.**
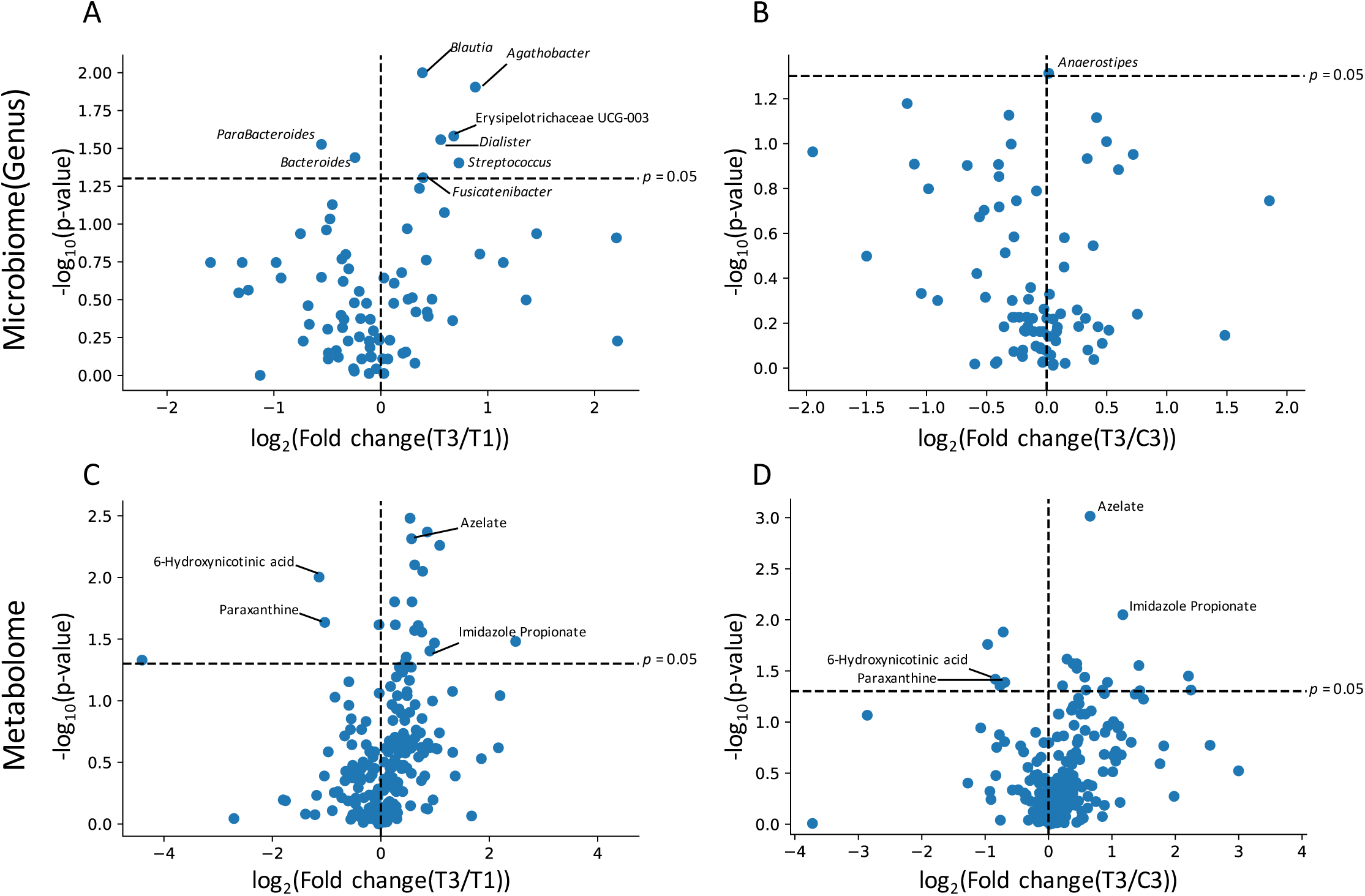
Effect of barely intake on intestinal microbiome and metabolome profiles. X axis indicates log fold change in mean value of corresponding genus/metabolite abundance after 4-week intervention relative to control time point. Y axis indicates logarithmic value of *p*-value. Each bacterium (A and B) and metabolite (C and D) were plotted. T1 (A and C) and C3 (B and D) were used as control time points.

### Characteristics of barley responders with glucose tolerance improvement

In a previous study, subjects were classified into responder and non-responder groups, based on phenotypic criteria to investigate the intestinal characteristics of subjects who showed improvement in glucose tolerance, due to barley consumption (5). We also adopted the criteria of glucose tolerance responders, reported in the study of Kovatcheva *et al*., revealing that only two or three subjects (subject 14 and 23 at T2, and subject 01, 02 and 06 at T3) were defined as glucose tolerance responders. Since analysis with small sample size could lead to incorrect results, we did not perform stratified analysis and instead performed correlation analysis. Specifically, for each subject, we defined the degree of glucose tolerance index improvement (referred to as glucose tolerance responder score), and performed correlation analysis between glucose tolerance responder score, and either baseline feature or change of feature by barley intake.

First, we analyzed glucose tolerance responder score and glucose tolerance index at baseline. We used blood glucose AUC/iAUC and insulin AUC/iAUC as the glucose tolerance indices. As a result, blood glucose AUC/iAUC and insulin AUC/iAUC improved in the individuals with high blood glucose AUC/iAUC and insulin AUC/iAUC at baseline, respectively (Fig 5). Interestingly, the improvement of blood glucose AUC/iAUC does not depend on insulin AUC/iAUC at baseline, and vice versa (Fig S2). This suggests that barley regulates glucose tolerance towards a healthier state and the improvement of the levels of blood glucose and insulin are independent of each other.

**Fig 5.**
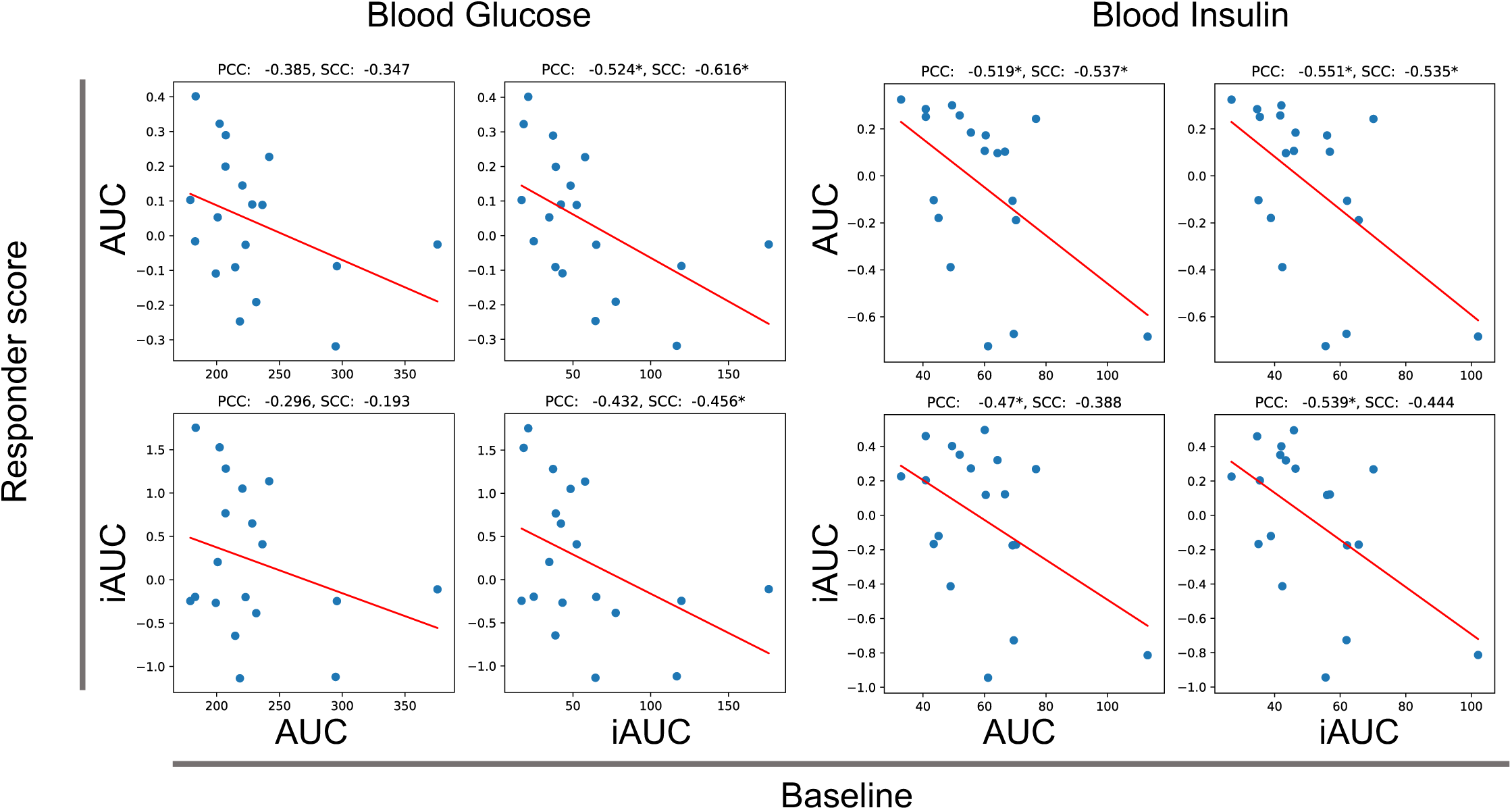
Improvement of glucose tolerance score by barley intake depends on glucose tolerance score at baseline. X axis indicates glucose tolerance index at baseline and Y axis indicates responder score. PCC:Pearson correlation coefficient, SCC: Spearman correlation coefficient, * *p* < 0.05, no correlation test

**Fig 6.**
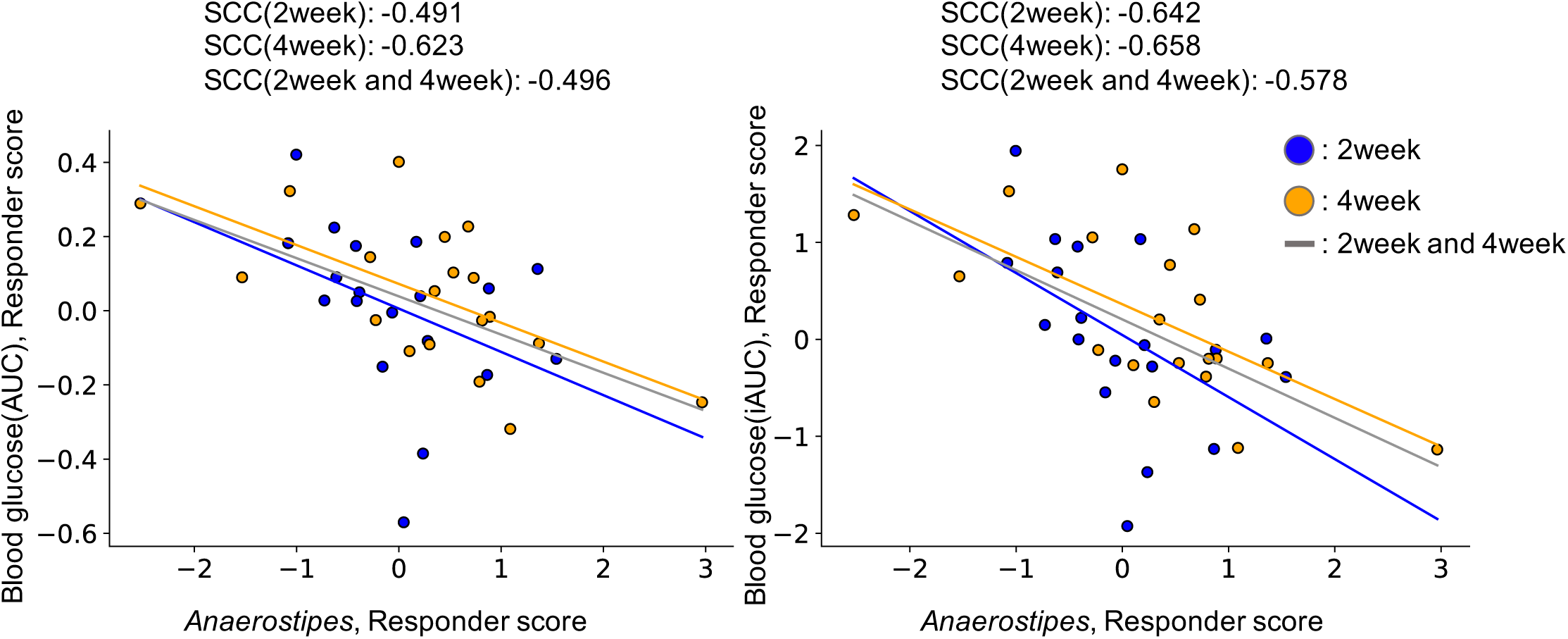
Increasing of *Anaerostipes* correlates with improved glucose tolerance. X axis indicates *Anaerostipes* responder score, and Y axis indicates blood glucose AUC/iAUC responder score.

Subsequently, we analyzed intestinal environmental changes that correlated with glucose tolerance index improvement by barley intake. As improvement of blood glucose AUC/iAUC did not depend on insulin AUC/iAUC at baseline, improvement analysis was performed independently. Although there were hundreds of intestinal bacteria and metabolites, few of them consistently correlated with blood glucose AUC/iAUC or insulin AUC/iAUC responder scores, across two time points (intake 2 weeks and 4 weeks). Only the increase of *Anaerostipes* by barley intake was consistently correlated with blood glucose AUC/iAUC improvement, across two time points (Fig 5; Table S4). Remarkably, the *Prevotella*/*Bacteroides* ratio was not correlated with glucose tolerance index improvement, although the *Prevotella*/*Bacteroides* ratio was previously reported to increase after barley consumption in responders (5). Blood glucose AUC/iAUC improvement correlated with some genera of *Prevotella* (*Prevotella* 2 and *Prevotella* 7) only at T3.

## Discussion

In our study, sodium, total protein and cholesterol levels in blood were significantly reduced, or had a declining tendency between T1 (before barley consumption) and T3 (4 weeks after barley consumption), which was consistent with previous reports (2, 9). Although there was a slight tendency, no significant difference was detected in the parameters between T3 and C3. Thus, we could not completely deny the possibility of a placebo effect, or an effect derived from the replacement of their regular staple food, with multi grain rice.

In the microbiome, microbial genera that belong to the family Lachnospiraceae, such as *Blautia, Agathobacter* and *Fusicatenibacter* showed significant increase in their relative abundance after barley consumption, compared to baseline (Fig 4A). Among those genera, *Blautia* and *Agathobacter* were previously shown to be increased by the intake of whole grain barley (10). We used pearled barley which lacks the bran and germ, and mainly consists of endosperm as the test food in our study. The endosperm and its contents contained in whole grain and pearled barley may be important factors for increasing these bacterial abundances. β-glucan is a soluble dietary fiber and a major component of endosperm, suggesting a strong effect of these fibers for the gut microbiota. It has been reported that *Blautia* abundance and visceral fat area is negatively correlated (11), and it is less prominent in the intestines of patients with diabetes or cirrhosis (12, 13). As such, *Blautia* is thought to be a putative biological marker for diseases related to metabolic syndrome such as obesity or diabetes. Although it is yet to be determined whether the increase in the abundance of *Blautia* has beneficial effects on these diseases, it could lead to improvement of their symptoms or conditions. *Agathobacter* is a bacterial genus where *Eubacterium rectale* has been reclassified and has been reported to be a dominant producer of butyric acid. Also, *Agathobacter* is known to be less prominent in the intestines of patients with ulcerative colitis, as compared to those of healthy individuals (14); however, the significant increase of butyric acid was not detected in this study. In a previous study, it was suggested that various subspecies exist within the *Agathobacter* genus, and subspecies that are dominant varies depending on the country (15). There is a possibility that the *Agathobacter* subspecies dominant in Japanese subjects’ intestines do not produce butyric acid, and it is also likely that although intestinal levels of butyric acid increase, fecal levels of butyric acid remain unchanged due to the produced butyric acid being absorbed in the small intestine (16).

In the metabolome profile, significant differences were observed in levels of azelate, paraxanthine, 6-hydroxynicotinic acid, and imidazole propionate in T3, compared to T1 and C3 (Fig 4C, D). As azelate is a common nutrient found in grains, including barley, it may not be increased due to the metabolism of intestinal microbiota. It has been reported that administration of azelate to mice improves the glucose tolerance, suggesting that it may be a factor contributing to the glucose tolerance improvement by barley consumption (17). Regarding imidazole propionate, a report has shown that it impairs insulin signaling via activating mechanistic targets of rapamycin complex 1 (18). Imidazole propionate produced in the gut reaches the liver through the portal vein, and impairs insulin signaling. Accordingly, its beneficial effect, when its amount is increased by barley consumption, is only worth considering when the imidazole propionate is excreted out of the intestine. By contrast, if intestinal imidazole propionate production is increased, there will possibly be a higher chance of absorption into the portal veins, which in turn leads to the deterioration of glucose tolerance. Even in high dietary fiber-containing barley, which is the substrate for SCFAs production, we did not detect any significant differences in SCFA levels at any time point.

During the study, subjects who showed improvement in the barley consumption-induced glucose tolerance had significantly high blood glucose iAUC prior to the barley intervention, suggesting that barley moderated the glucose tolerance towards a healthier state. Subsequently, we explored bacteria and metabolites that correlated with blood glucose AUC/iAUC and insulin AUC/iAUC responder scores. Previous study showed that barley consumption increases *Prevotella/Bacteroides* ratio in barley-consuming healthy Swedish subjects with improved glucose tolerance (5). In this study, increasing of *Prevotella*/*Bacteroides* ratio does not correlated with blood AUC/iAUC improvement but *Prevotella* 2 and *Prevotella* 7 correlated with blood glucose AUC/iAUC improvement. It is assumed that i) increasing of *Prevotella*/*Bacteroides* ratio by intake of barley is not maintained for a long time (intervention period was 3 days in previous study and 14 days and 28 days in this study) ii) *Prevotella* functions are dissimilar by various strains of *Prevotella* in multiple countries (7). *Anaerostipes* was correlated with blood glucose AUC/iAUC improvement consistently. Abundance of *Anaerostipes* was higher in the low glycemic index diet intake group (19). In addition, in two independent studies, it was found that the presence of *Anaerostipes hadrus* that encodes a composite inositol catabolism-butyrate biosynthesis pathway resulted in lower host metabolic disease risk (20). Increase of *Anaerostipes* by barley intake may be related to the improvement of glucose tolerance; however, Zeevi *et al*., described that there are structural variations in the *Anaerostipes hadrus* genome. In 16S rRNA gene analysis, it is unclear whether *Anaerostipes* encodes composite inositol catabolism-butyrate biosynthesis pathway gene, so additional testing using shotgun metagenomics may be necessary.

This study has a few limitations. First, the control food also contained dietary fibers. Dietary fiber itself acts as a prebiotic; also known as microbiota-accessible carbohydrates (MACs), to produce short-chain fatty acids (SCFAs) by gut microbiota (21), thus having potentially similar effects as barley. Therefore, it may be difficult to see the impact of test food clearly, as compared with control food intake. Second, intestinal microbes and metabolites are complex parameters which contain many different bacterial genera and metabolites. FDR correction is necessary in statistical hypothetical tests; however, as many items were comprehensively observed, the FDR correction was strict, increasing the difficulty in detecting the true significant differences. Therefore, validation tests using models such as murine models, should be considered to confirm these findings.

This randomized controlled study provided novel insights into the effect of barley on the intestinal environment and blood parameters in a Japanese population. Barley intake increased some intestinal bacteria such as *Blautia* and *Agathobacter*, and metabolites such as azelate. In addition, the improvement of glucose tolerance due to the effect of barley intake was dependent on the baseline glucose tolerance, indicating that an increase in the gut *Anaerostipes* is possibly involved in its improvement. In this study, a metabologenomic approach, together with a randomized, double blind, placebo-controlled crossover clinical trial, uncovered the impact of diet on the intestinal environment, and shed light on stratified health care considering the intestinal environment.

## Materials and Methods

### Ethics approval

The human rights of the subjects who participated in this study were protected at all times, and the study observed the Helsinki Declaration and Ethical Guidelines on Epidemiological Research in Japan referring to cases concerning standards for clinical trials of drugs. Informed consents were obtained from all individual participants and preserved in the text. This trial was conducted with approval of the clinical trial ethics review committee of Chiyoda Paramedical Care Clinic (UMIN-CTR, Trial number: UMIN000023675).

### Trial design and recruitment

In this study, a randomized double-blind controlled trial in Japanese participants were performed for 3 months. The study included 4-week dietary intervention periods (Period 1 and 2), comprising a test food, 32% barley-blended multi grain rice, and control food, barley-free multi grain rice, in random order and separated by a 4-week washout period (Washout) (Fig 1, Fig S3, Table S5). Barley was shaped by cutting and pearling to mimic rice. During the dietary intervention periods, subjects were instructed to substitute the staple food in their diet with the test or control food twice a day. The test food and control food were preliminary processed to avoid subjects to identify the difference visually. The trial was initiated in July 2016 and completed in December 2016.

During the trial, fecal samples were collected at baseline (T1 and C1, T = test food and C = control food), 2 weeks (T2 and C2), and 4 weeks (T3 and C3) of the dietary intervention periods. The collected stool samples were frozen at -20°C until processing. In addition, clinical blood tests and oral glucose tolerance tests were performed following a 12-h fasting at the same time points. In the clinical blood test, total protein, albumin, aspartate aminotransferase, alanine transaminase, lactate dehydrogenase, total bilirubin, alkaline phosphatase, γ-glutamyl transpeptidase, creatine phosphate enzyme, urea nitrogen, creatinine, uric acid, sodium, chlorine, potassium, calcium, total cholesterol, low-density lipoprotein cholesterol, high-density lipoprotein cholesterol, neutral fat, glucose, insulin, hemoglobin A_1c_, white blood cell, red blood cell, hemoglobin, hematocrit, and platelet were measured. For oral glucose tolerance test, blood samples were taken at 30, 60, 90 and 120 min after oral administration of 75 g glucose for glucose and insulin level measurements. The AUC and iAUC were calculated using the trapezoid model.

In total, 48 participants were recruited in this study. The participants fulfilled the following criteria: aged between 50 and 69 years old, body mass index of 18–25 kg/cm^2^, and fasting plasma glucose level under 109 mg/dL. Detailed inclusion/exclusion criteria are shown in Table S6. Prior to the main trial, the blood tests were conducted. Based on the preliminary blood test and male-female ratio, 24 subjects were selected for the main trial. All 24 subjects completed the trial; however, five subjects were excluded from the analysis for following reasons: incomplete blood sampling (subject 11), incomplete fecal sampling (subject 13), and consumption of prohibited food such as fermented products during the trial (subjects 08, 18, and 22). The primary outcome was blood glucose and insulin AUC after 2 weeks and 4 weeks of test food consumption. In addition, the key secondary outcome was blood glucose and insulin iAUC, fasting blood glucose and insulin, and stool frequency.

### Trial intervention: randomization and blinding

Randomization in this trial was performed using the blocked stratified randomization method with the subject assignment manager. First, 24 subjects who passed the inclusion criteria were assigned to two groups (group A and B) by stratification, taking into consideration the age and male-female ratio immediately before the trial period.

Subsequently, the symbols “A” or “B”, representing the test food and control food, respectively, were randomly assigned to each group of subjects. Following, the test food assignment table with the test food symbol and the subject identification code was prepared. Immediately after the assignment to the test food according to the assignment table, the table was sealed and kept tightly concealed by the subject assignment manager. The table was disclosed to the test analyst, investigator, and test sharing doctor after data fixation.

### DNA extraction and 16S rRNA gene-based microbiome analysis

DNA extraction from fecal samples was performed as previously reported (22). After extraction, the V1-V2 variable region of the 16S rRNA gene was amplified using bacterial universal primers 27F-mod (5′-AGRGTTTGATYMTGGCTCAG-3′) and 338R (5′-TGCTGCCTCCCGTAGGAGT-3′) with Tks Gflex DNA Polymerase (Takara Bio Inc., Japan) (23). The amplicon DNA was sequenced using MiSeq (Illumina, USA) according to the manufacturer’s protocol. All 16S rRNA amplicon sequence files generated in this study are available in DRA of DDBJ (DRA accession number: DRA009319).

### Metabolite extraction and CE-TOFMS-based metabolome analysis

Extraction of metabolites from fecal samples was performed as previously reported (24). Briefly, the samples were initially lyophilized using the VD-800R lyophilizer (TAITEC) for at least 24 h. Freeze-dried feces were disrupted with 3.0 mm zirconia beads by vigorous shaking (1,500 rpm for 10 min) using the Shake Master neo (Biomedical Science). Next, 500 µL of methanol, including the internal standards (20 µM each of methionine sulfone and D-camphor-10-sulfonic acid (CSA)), was added to 10 mg of disrupted feces. Samples were further disrupted with 0.1 mm zirconia/silica beads by vigorous shaking (1,500 rpm for 5 min), then 200 µL of ultrapure water and 500 µL of chloroform were added before centrifugation at 4,600×g for 15 min at 20°C. Subsequently, 150 µL of the aqueous layer was transferred to a centrifugal filter tube (Ultrafree MC-PLHCC 250/pk for Metabolome Analysis, Human Metabolome Technologies) to remove protein and lipid molecules. The filtrate was concentrated by centrifugation and dissolved in 50 µL of ultrapure water immediately before coupling capillary electrophoresis with electrospray ionization time-of-flight mass spectrometry (CE-TOFMS) analysis. The obtained metabolome data are available in Tables S7 and S8.

### Bioinformatics and statistical analysis

For 16S rRNA gene analysis, QIIME2 (version 2019.10) was used (25). In the analytical pipeline, sequence data were processed by using the DADA2 pipeline for quality filtering and denoising (options: --p-trim-left-f 20 --p-trim-left-r 19 --p-trunc-len-f 240 --p-trunc-len-r 140) (26). The filtered output sequences were assigned to taxa by using the “qiime feature-classifier classify-sklearn” command with the default parameters. Silva SSU Ref Nr 99 (version 132) was used as a reference database for taxonomy assignment (27). The microbiome data are available in Table S9

In statistical analysis to compare primary and secondary outcomes, we used paired *t*-test and paired Cohen’s d using R package effsize. Other all statistical analyses were performed using Python scripts. Multidimensional scaling was performed using unweighted UniFrac distance and Spearman correlation distance calculated from the data of microbiome OTUs and metabolite relative area, respectively. In addition, intra-distance was defined as a set of all-to-all distance in all samples from the same subjects, excluding the distance between the same samples, for all individuals, and inter-distance as all-to-all distances in all T1 samples, excluding the distance between the same samples. For the pairwise comparison in relative abundance of intestinal bacteria and relative area of intestinal metabolite, Wilcoxon signed-rank test with Benjamini-Hochberg false discovery rate correction was used. During the comparison, bacteria with relative abundance below 0.001 were excluded. Further, Wilcoxon rank sum test with Benjamini-Hochberg false discovery rate correction was used for comparison between responder and non-responder groups.

### Defining responders with specific response

We also adopted criteria of glucose tolerance responders reported in the study of Kovatcheva *et al*. (insert date of study), in which responders must fulfill the following criteria: (1) comparing T2 and C2 or T3 and C3, blood glucose iAUC (0–90 min) decreased by at least 25%; (2) comparing T2 and C2 or T3 and C3, blood glucose AUC (0– 90 min) decreased; (3) comparing T2 and C2 or T3 and C3, insulin iAUC (0–90 min) decreased by at least 15%. In this study, the test food effect size was defined as the responder score and used to evaluate whether effects depended on individual basal characteristics. The response score was calculated with the following equation:

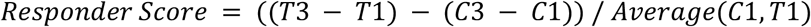

## Data Availability

All 16S rRNA amplicon sequence files generated in this study are available in DRA of DDBJ (DRA accession number: DRA009319)

## Acknowledgements

The super-computing resource was provided by the Human Genome Center, the Institute of Medical Science, the University of Tokyo. We would like to thank Hakubaku Co., Ltd. and CPCC Co., Ltd. to conduct the clinical trial. Additionally, we would like to thank the Metabologenomics, Inc. staff who participated in discussion of this research.

## Author contributions

Conceptualization, T.M, T.Y, Toshiki K and S.F; Data Curation, Y.N; Formal Analysis, Y.N; Investigation, Y.M and M.I; Methodology, Y.G and S.M; Project Administration, Toshiki K, and S.F; Resources, Y.G and S.M; Supervision, Toshiki K and S.F; Visualization, Y.N and T.Y; Writing – Original Draft, Y.N; Writing – Review & Editing, Y.G, Y.N, T.N, R.N, Toru K, T.M, T.Y, Toshiki K, and S.F

## Supplemental information titles and legends

**Fig S1.**
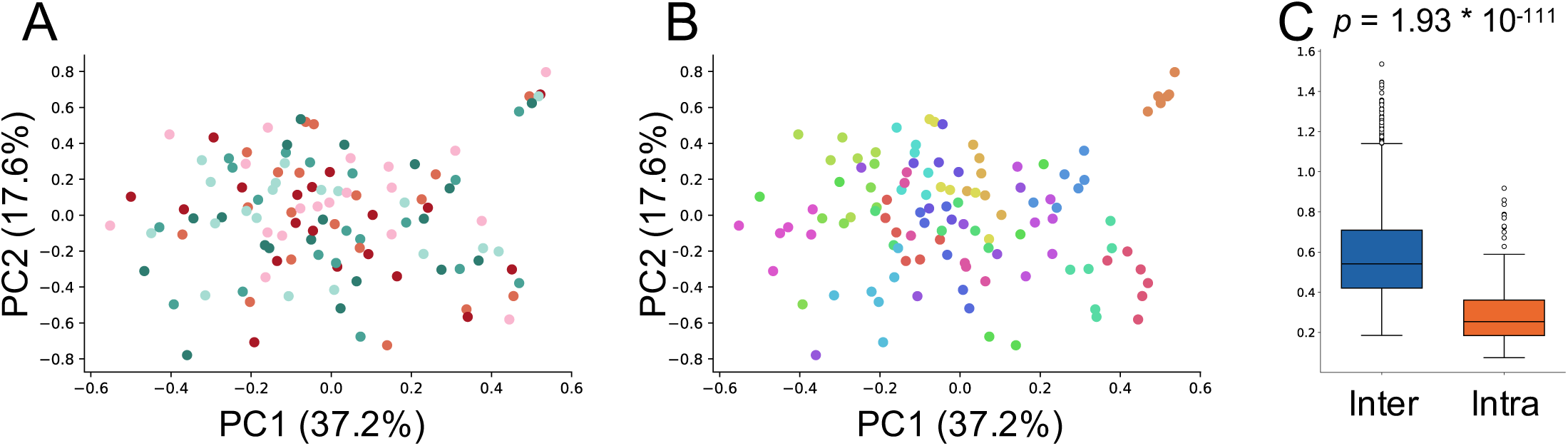
Individual differences were larger than the influence of barley or control food consumption on microbiome (weighted UniFrac distance) (A, B) Scatter plots showing results of multidimensional scaling using beta diversity (weighted UniFrac distance) calculated from microbiome profile. Plots were color-coded by subject (A) or time point (B). (C) Box plot representing the distribution of unweighted UniFrac distance between samples from different subjects at the same time point (inter-individual, represented in blue) and the distance between samples from the same subject (intra-individual, represented in orange).

**Fig S2.**
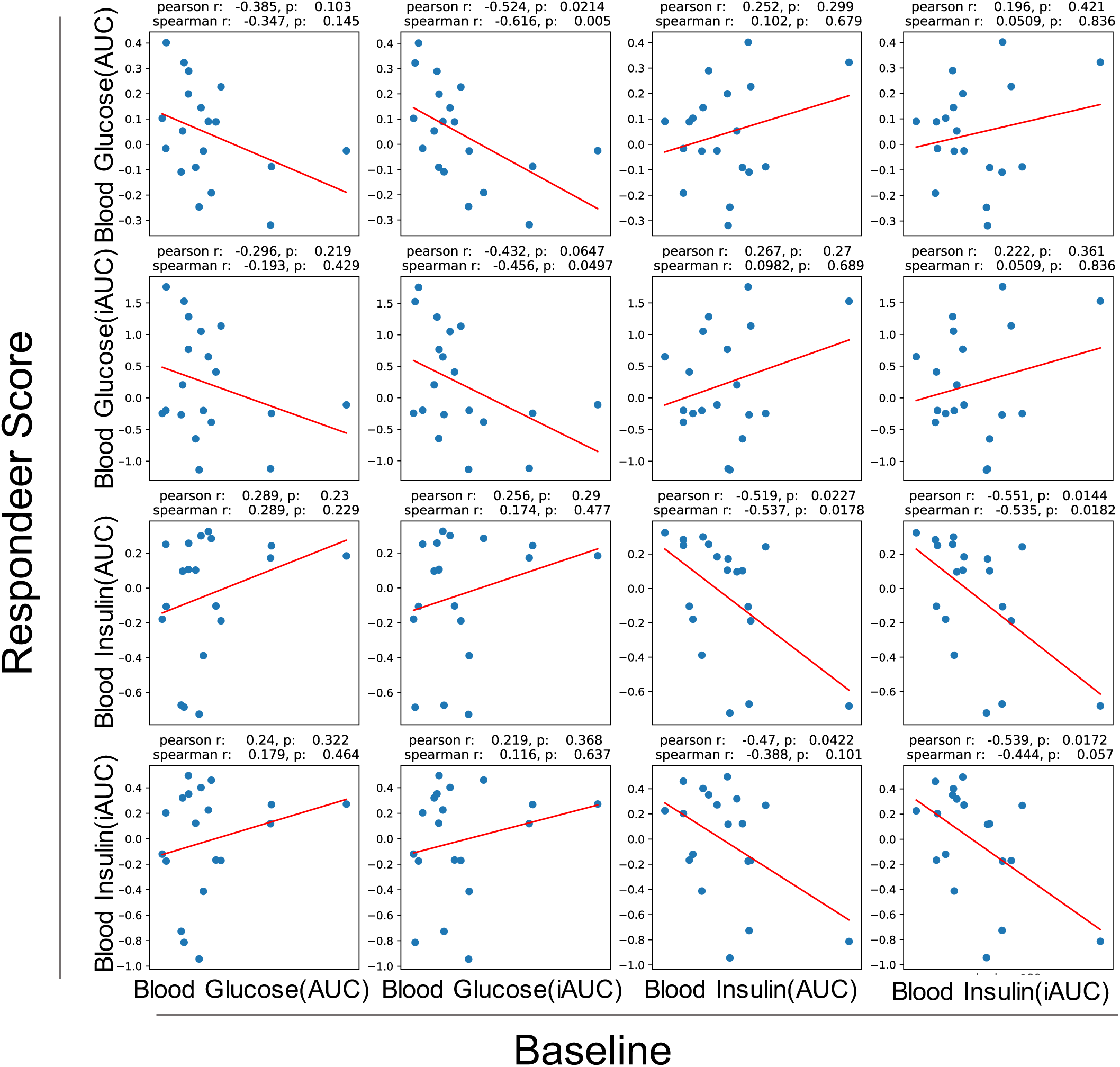
Improvement of blood glucose AUC/iAUC by barley intake does not depends on blood insulin AUC/iAUC at baseline and vice versa. X axis indicates glucose tolerance index at baseline and Y axis indicates degree of improvement of glucose tolerance index. p-value is no correlation test.

**Fig S3.**
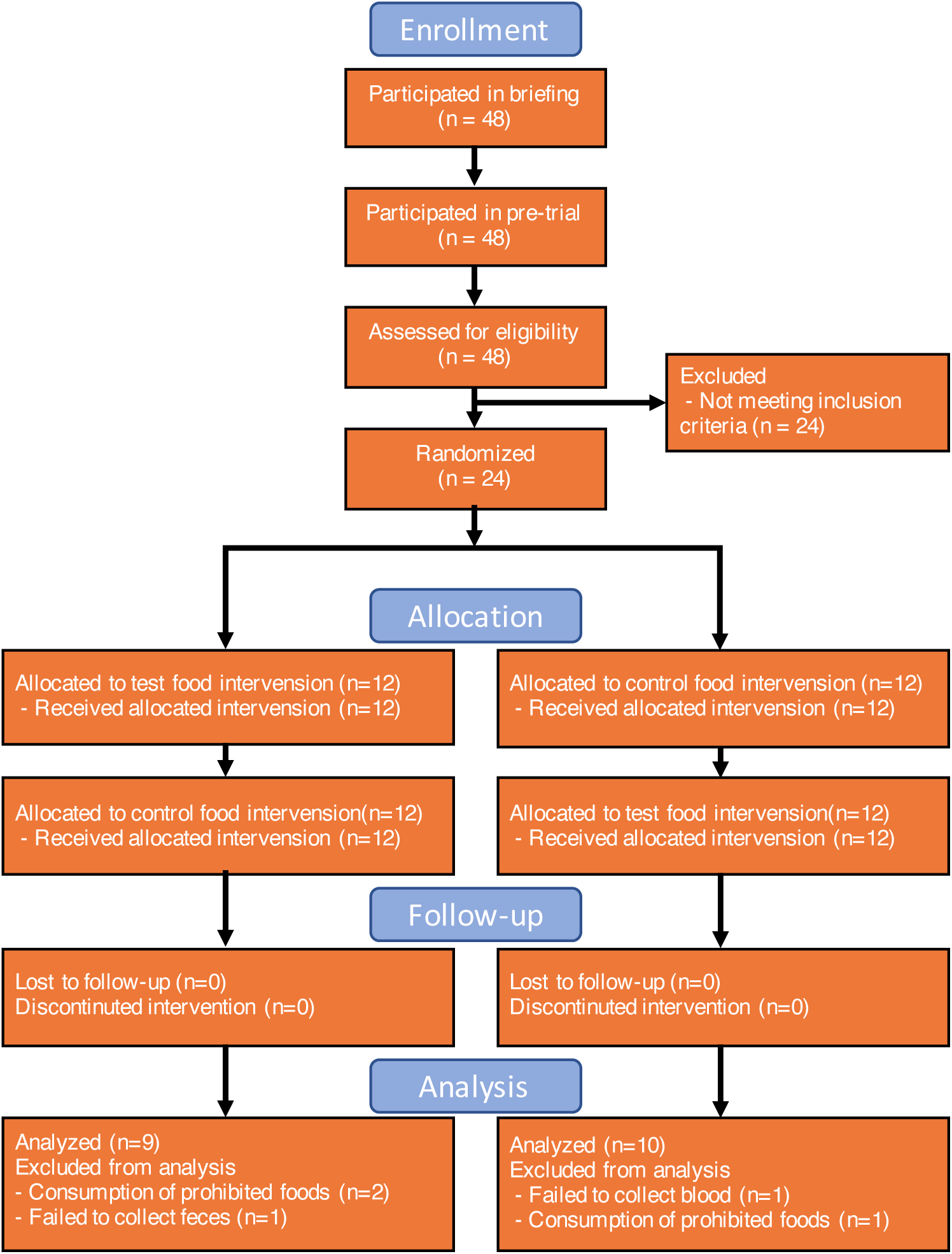
Flow diagram of the double-blind, placebo-controlled crossover study.

**Table S1. Difference in primary/secondary outcome in baseline**.

**Table S2. Difference in gut microbiota between timepoints**

**Table S3. Difference in gut metabolite between timepoints**

**Table S4. Significantly correlated bacteria and metabolites with blood glucose responder score**

**Table S5. CONSORT checklist**

**Table S6. Key inclusion/exclusion criteria**

**Table S7. Relative area of metabolome in feces**

**Table S8. Amount of metabolome in feces**

**Table S9. Microbiome phylogenetic composition**

